# Active commuting, anxiety symptoms and mental wellbeing: a dose-response study

**DOI:** 10.64898/2026.06.12.26355515

**Authors:** Laura Joensuu, Juuso J. Jussila, Timo Lanki, Pekka Tiittanen, Tytti P. Pasanen, Ulf Ekelund, Jaana I. Halonen

**Author notes:** Correspondence to: Laura Joensuu, Finnish Institute for Health and Welfare, Finland.

## Abstract

Climate change draws attention to the planetary health perspective in sport and exercise sciences, that is, to physical activity that supports both human wellbeing and environmental sustainability. Active commuting is a sustainable form of physical activity with well-established somatic health benefits. However, more knowledge is needed on its relationship with mental health. We examined dose–response associations between active commuting, anxiety symptoms, and mental wellbeing among Finnish adults, and whether green commuting environment moderates these relationships. We used data from the cross-sectional Environment and Health Survey collected in June-September 2023 in the ten largest cities in Finland. Employed participants with data on anxiety symptoms (Generalized Anxiety Disorder-7, GAD-7), mental wellbeing (World Health Organization-Five Well-Being Index, WHO-5), commuting profile over a year (mode, frequency, distance, and perceived greenness along the commute route), and sociodemographic and lifestyle factors were included (n=1,672; mean age 45.3 years; 53.8% women). Active commuting was defined as travelling the entire commute by walking or cycling (including e-biking) that was converted into approximated annual km/week and MET-h/week. We used linear and logistic regression with restricted cubic splines to evaluate dose-response associations, adjusted for key covariates. The role of perceived greenness was tested using an active commuting × commute greenness interaction term. We found no dose-response relationships between active commuting and anxiety symptoms or mental wellbeing in any of the models. No effect modification by commute greenness was observed. More research on how active commuting may support planetary health from a mental health perspective is needed.

## Introduction

Planetary health is a framework that recognizes the interdependence between human wellbeing and the natural systems that sustain life.^1^ It therefore draws attention to lifestyle behaviors that support both human health and environmental sustainability.^2^ For example, transitions toward more plant-based diets have gained significant attention within this framework,^3^ including their incorporation into Nordic nutritional recommendations.^4^ However, physical activity has not yet received a similar status despite calls for integrating sustainability perspectives also into physical activity and sedentary behavior guidelines.^5^

Transportation remains a major source of greenhouse gas emissions,^6^ and active travel (i.e., walking and cycling for transportation) is considered a physical activity domain with considerable potential to support both human health and the health of the Earth’s natural systems.^2^ Commuting is one of the primary reasons for daily transportation,^7^ and changes to active modes have shown several environmental co-benefits, such as reduced greenhouse gas and air pollution emissions.^8^ Regular active commuting has also well-established somatic health benefits, particularly related to cardiometabolic health, obesity, physical fitness and longevity.^9–12^ However, its positive relationship with mental health is less evident.

Previous studies have reported small positive, null, or even adverse associations between active commuting and mental health.^13–29^ Some prior findings have indicated that the mental health outcomes of active commuting may be dose-specific. For example, a study of Finnish adults found that high volumes of active commuting (30 minutes or more a day) were associated with higher odds of depressive symptoms, while no associations were observed for lower volumes.^29^ Less is known if a similar pattern exists for other mental health outcomes such as anxiety and mental wellbeing.

Furthermore, environmental features such as greenness along commuting routes may facilitate positive affect and modify mental health outcomes, offering alternative explanation for previous inconsistent findings.^19,30^ Greenness has been found to provide greater benefits for mood and affective outcomes of physical activity, in comparison to urban built environments.^31^ Relevant frameworks include Attention Restoration Theory,^32^ and the Stress Recovery Theory,^33^ which suggest that exposure to natural environments may enhance cognitive functioning and parasympathetic activity, leading to restored attentional capacity and relieved stress.^34^ Potential psychophysiological pathways include improvements in executive function, heart-rate variability, and brain wave activity and connectivity.^34–36^

From the planetary health perspective, it remains viable to study how active commuting is associated with mental health and factors which potentially moderate this relationship. In this study, we examined the dose-response relationship between active commuting, anxiety symptoms, and mental wellbeing among employed adults in Finland. In addition, we tested whether green commuting environments modify these associations.

## Methods

### Study design and population

We used data from the cross-sectional Environment and Health Survey collected by the Finnish Institute for Health and Welfare (THL) in June-September 2023. The questionnaire was mailed to a random population-based sample of over 20-year-old inhabitants living in the ten largest cities in Finland (N=20,000), and it was available in three language versions (Finnish, Swedish and English).^37^ The questionnaire included items related to individual-level socioeconomic factors, health and wellbeing, lifestyle, living environment, and climate attitudes and beliefs. Response rate was 34% and a total of 5,879 responses were received (age range 20–98 years). We used a complete-case approach and excluded participants who were retired, on a family leave, stay at home parents, students, unemployed or laid off (n=3,228), had >20km one-way commute (n=585, the cut-off was based on national survey data of feasible active commuting distances)^7^, had used medication during past 30 days for anxiety (n=155) and/or depression (n=88), and had missing data on exposures (n=11), outcomes (n=39), or covariates (n=93). Additionally, those who constituted a limited demographic group or were outliers limiting modelling were excluded (non-binary gender, n=6; BMI>60, n=2). Final analytical sample included 1,672 participants (Figure 1).

**Figure 1.**
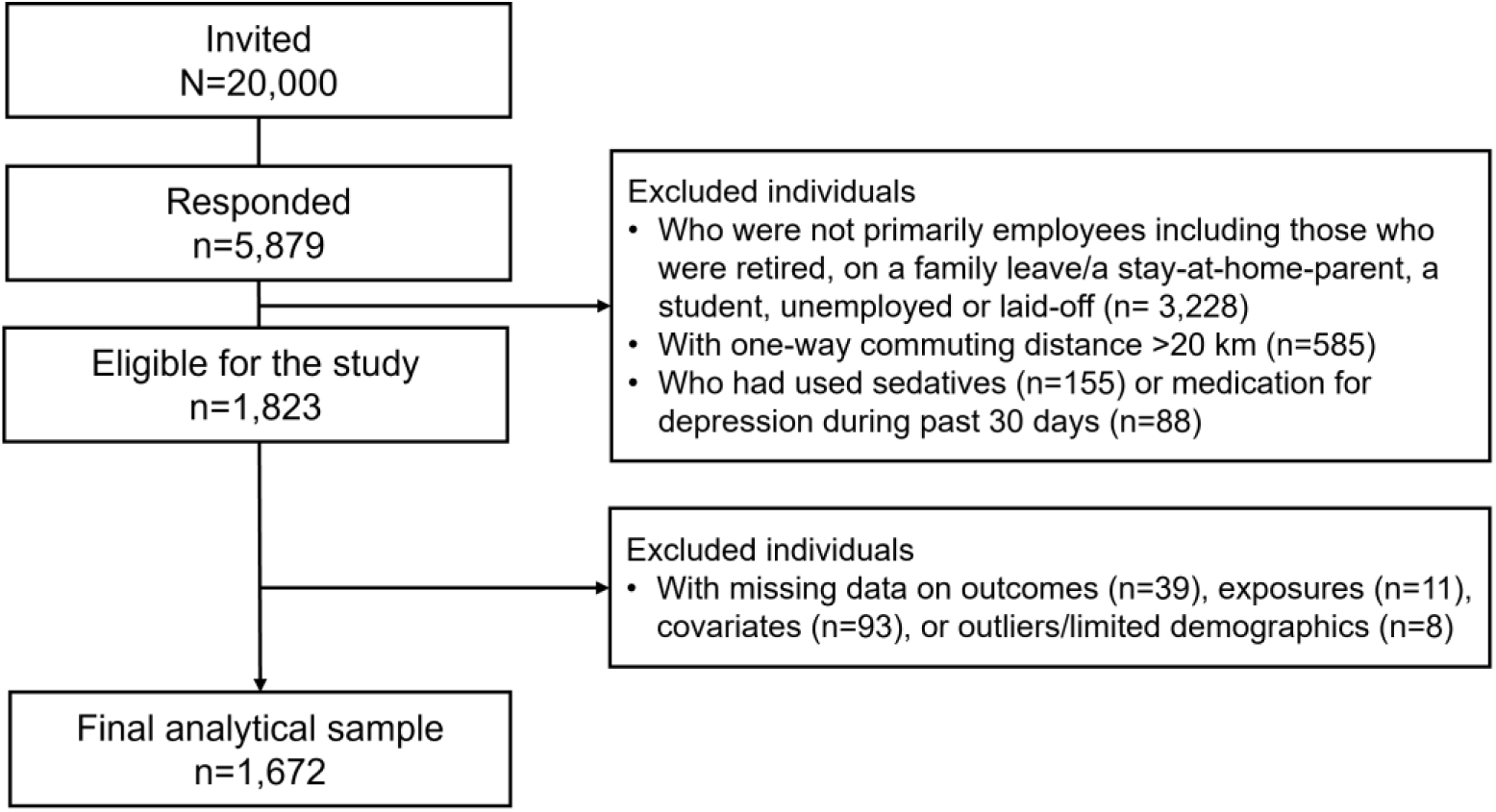
Flow chart of participant selection.

The data collection was approved by THL Ethics Working group (THL/706/6.02.01/2023) and conducted in accordance with the Declaration of Helsinki. Respondents were informed about the use and handling of data in the covering letter, and they accepted the use of data for scientific research by returning the survey. The reporting in this study adheres to the STROBE guidelines.^38^

### Exposures

We used self-reported commuting mode, frequency, and distance to derive active commuting variables. Summertime and wintertime commuting behaviours were reported separately. Participants reported whether they 1) walked the whole way, 2) cycled the whole way, 3) cycled the whole way with an e-bike, 4) used public transport with ≥1 km walking or cycling, 5) used public transport with <1 km walking or cycling, or 6) used private car (as a passenger or driver) or other motorized vehicle (e.g., motorcycle), including the weekly frequency for using each commuting mode. The reported frequencies were transformed as follows: “never” (0 days), “1–2 days per week” (1.5 days), “3–4 days per week” (3.5), and “5 days of more often” (5 days) and were summed across walking and cycling (including e-biking). A maximum number of seven commuting days per week was applied if summed frequencies were higher. Participants additionally reported their one-way commuting distance in kilometres, which was multiplied by two for daily commuting distance.

We derived the approximated annual active *km/week* commuting by multiplying the number of active commuting days (walking and cycling) by the daily commuting distance, calculated separately for cold and warm season. We then derived a weighted average based on seven months (April–October) during which roads are ice-free, called here for simplicity summer months, and five winter months (November–March), reflecting typical Finnish climate conditions. Kilometres were selected as the exposure metric because they are commonly used in population health impact tools, such as the World Health Organization’s Health Economic Assessment Tool (HEAT).^39^

To align with the evidence base used for physical activity guidelines, we additionally calculated a *MET-h/week* variable. We used metabolic equivalents of task (METs) reported in the Global Physical Activity Questionnaire Analysis Guide for active travel (walking and cycling: 4 METs),^40^ and corresponding approximated walking and cycling speeds (km/h) based on the latest release of the Compendium of Physical Activities (walking: 5 km/h, cycling: 15 km/h).^41^ We used the following formulas for the calculation:

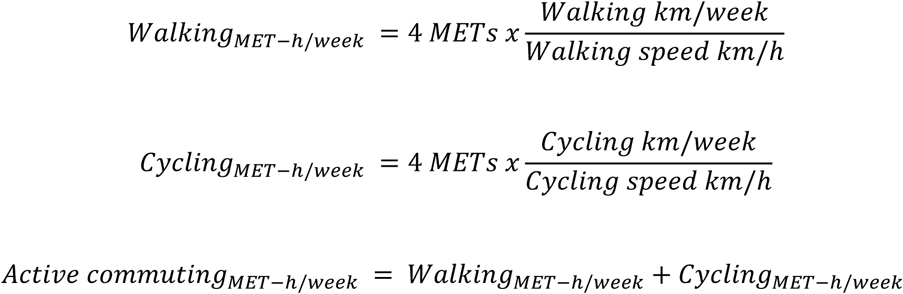

We calculated summertime-only km/week and MET-h/week variables for sensitivity analyses.

### Outcomes

*Anxiety symptoms* were surveyed using the Generalized Anxiety Disorder-7 (GAD-7) questionnaire.^42^ The participants reported how often the following problems have bothered them during the past two weeks: 1) feeling nervous, anxious, or on edge, 2) not being able to stop or control worrying, 3) worrying too much about different things, 4) having trouble relaxing, 5) being so restless that it is hard to sit still, 6) becoming easily annoyed or irritable, and 7) feeling afraid as if something awful might happen.

The answers on a 4-point Likert scale (from 0 = not at all to 3 = nearly every day) were summed and formed a score between 0 and 21. The score was divided into four categories following the recommendations of the scale developers: minimal (0–4), mild (5–9), moderate (10–14) and severe (15–21) anxiety.^42^ Data was skewed and there were limited number of moderate and severe symptoms. Therefore, a binary variable (0 = minimal, 1 = at least mild anxiety symptoms) was used in the main analyses. All symptom levels (mild, moderate and severe) are associated with lower health-related quality compared with minimal symptoms.^42^ The categorical variable was used for descriptives and the continuous for correlation analysis. The internal consistency of the GAD-7 in this data was checked and considered good (Cronbach’s α=0.86).

*Mental wellbeing* was assessed using the World Health Organization-Five Well-Being Index (WHO-5), a widely used measure with well-documented psychometric properties.^43^ The following was requested: “Please choose, for each of the five statements, the option that best describes how you have felt during the past two weeks.” 1) I have felt cheerful and in good spirits, 2) I have felt calm and relaxed, 3) I have felt active and energetic, when waking up, 4) I have felt fresh and rested, and 5) my daily life has been filled with many things that interest me. The answers on a 6-point Likert scale (from 0 = not at all to 5 = all the time) were summed and multiplied by 4, following the WHO guidelines.^43^ This formed a score between 0 and 100 which was used as a continuous variable in the analyses. The internal consistency of the WHO-5 in this data was checked and considered good (Cronbach’s α=0.87).

### Moderator

Perceived commute greenness was assessed using a question “How much green space is there along your commuting route?” 1) none or a very small part of the journey, 2) about one quarter of the journey, 3) about half of the journey, 4) about three quarters of the journey, and 5) the entire or almost the entire journey. A binary moderator variable was formed based on the distribution of responses indicating less than 50 % (answers 1–2) or 50 % or more of green areas (answers 3–5).

### Covariates

Covariates were self-reported and selected based on previous literature.^14,22^ Of basic demographics, we used age and gender. Educational attainment was characterised as a binary variable indicating completion of either basic/secondary or tertiary education, following the International Standard Classification of Education (ISCED) standards. Basic/secondary education corresponds to ISCED levels 1–5 and tertiary education ISCED levels 6–8.^44^ We used a binary variable for income satisfaction (no difficulties or difficulties in making ends meet) and relationship status (having or not having a partner).

Smoking was evaluated as smoked cigarettes per day on a six-item scale and transformed into a binary variable of no smoking/current smoker due to the low number of frequent smokers. Alcohol consumption was assessed with a five-item frequency question, where the two upper categories were combined due to the low prevalence of frequent alcohol use in the cohort: “never”, “once per month or less”, “2–4 times per month”, and “2–3 times per week or more”. Alcohol consumption quantity was also requested but excluded from further analyses because 11% of the data was missing.

Leisure-time physical activity (LTPA) was assessed with a binary variable indicating regular physical activity during leisure-time or not. Occupational physical strain included four items, where the two most strenuous categories were combined: “mostly sedentary”, “walking, no heavy lifting”, and “physically demanding”. Body mass index (BMI) was based on self-reported height and weight (weight kg / height m^2^) and used as a continuous variable.

### Statistical analysis

We calculated standard descriptives for the study variables and additionally examined Spearman correlations between the active commuting exposures, continuous outcomes, and categorical commute greenness and covariates. We also described primary commuting modes during wintertime and summertime separately and illustrated transitions between seasonal commuting modes using reported commuting frequencies and alluvial plots.

We analyzed the associations between active commuting and mental health with regression analyses separately for anxiety symptoms and mental wellbeing outcomes. In *a priori* power calculations, a sufficient sample size was N>868, with ɑ-level set to 0.05, power 0.80, 11 predictors, and a small effect size (≤0.14). Previous studies have indicated that active commuting may have non-linear associations with mental health,^29^ and thus we applied analytical methods that allow examination of non-linear dose-response patterns. We used logistic regression for the binary anxiety symptoms and linear regression for the continuous mental wellbeing score with restricted cubic splines. We employed the Akaike information criterion (AIC) and overfitting indices to select the optimal number of knots. Three knots (placed at 5%, 50% and 95%) showed the best fit and the lowest overfitting indices and were used throughout the analyses.

For both regression analyses, the main model included the active commuting variable (km/week or MET-h/week), age, gender, commute greenness, educational attainment, income satisfaction, relationship status, smoking, alcohol use, LTPA, and occupational physical strain. In sensitivity analyses, we further adjusted for BMI, as it may act as a confounder, mediator, or collider in the model. We also provide the minimally adjusted associations (only with age and gender) in the Supplement. We tested moderation by adding a multiplicative active commuting × commute greenness interaction term. While the whole-year commuting patterns were used in the main analyses, summertime commuting (indicating a closer relation to outcomes experienced during the past two weeks) was used in sensitivity analyses.

We additionally evaluated face validity of the commute greenness variable against participants’ satisfaction with residential area characteristics. In the survey, this was requested using 14 items reflecting satisfaction ranging from green and blue spaces to outdoor air quality and noise level. Higher perceived commute greenness was associated with better satisfaction with residential area characteristics, such as green areas and overall greenery (Supplemental Figure 1).

All analyses were conducted with R (version 2026.01.2). Statistical significance level was set to p<0.05 throughout the study.

## Results

Characteristics of the study sample are presented in Table 1. Participants were on average 45.3 years old, most of them had a tertiary education (63%), were comfortable with their income (91%), had a partner (78%), were non-smokers (86%), were regularly physically active during leisure-time (88%), and were mostly sedentary in their work (61%). For alcohol consumption, the highest prevalence was in the 2–3 times per month category (39%). The majority (52%) did not commute actively to work. Most participants experienced minimal anxiety symptoms (83%), and on average they had a good mental wellbeing score (65.3). A Spearman correlation heatmap between active commuting and other variables is presented in Supplemental Figure 2. Higher active commuting (MET-h/week) correlated with higher anxiety symptoms (ρ=0.05, p=0.045), but no other correlations between active commuting and mental health were observed. Greater commuting greenness was correlated with lower anxiety symptoms (ρ=-0.06, p=0.022), better mental wellbeing (ρ=0.11, p<0.001), and higher active commuting (km/week ρ=0.18, p<0.001 and MET-h/week ρ=0.17, p<0.001).

**Table 1.** Descriptives statistics of the study sample.

| Characteristics | All | Women | Men |
| --- | --- | --- | --- |
| **N (%)** | 1672 (100%) | 899 (53.8%) | 773 (46.2%) |
| **Age (years)** | 45.3 (12.9) | 45.2 (13.3) | 45.3 (12.6) |
| **Educational attainment** |  |  |  |
| Basic/Secondary | 621 (37.1%) | 317 (35.3%) | 304 (39.3%) |
| Tertiary | 1051 (62.9%) | 582 (64.7%) | 469 (60.7%) |
| **Income satisfaction** |  |  |  |
| No difficulties in making ends meet | 1526 (91.3%) | 800 (89.0%) | 726 (93.9%) |
| Difficulties in making ends meet | 146 (8.7%) | 99 (11.0%) | 47 (6.1%) |
| **Has a partner** | 1305 (78.1%) | 685 (76.2%) | 620 (80.2%) |
| **Smoking** |  |  |  |
| No smoking | 1444 (86.4%) | 785 (87.1%) | 661 (85.5%) |
| Current smoker | 228 (13.6%) | 116 (12.9%) | 112 (14.5%) |
| **Alcohol use** |  |  |  |
| Never | 177 (10.6%) | 111 (12.3%) | 66 (8.5%) |
| Once per month or less | 437 (26.1%) | 291 (32.4%) | 146 (18.9%) |
| 2–4 times per month | 643 (38.5%) | 333 (37.0%) | 310 (40.1%) |
| 2–3 times per week or more | 415 (24.8%) | 164 (18.2%) | 251 (32.5%) |
| **Body mass index (kg/m<sup>2</sup>)** | 26.1 (4.7) | 25.7 (5.1) | 26.5 (4.1) |
| **Regular leisure-time physical activity** | 1468 (87.8%) | 798 (88.8%) | 670 (86.7%) |
| **Occupational physical strain** |  |  |  |
| Mostly sedentary | 1015 (60.7%) | 513 (57.1%) | 502 (64.9%) |
| Walking, no heavy lifting | 321 (19.2%) | 213 (23.7%) | 108 (14.0%) |
| Physically demanding | 336 (20.1%) | 172 (19.2%) | 163 (21.1%) |
| **Commuting actively over the year** |  |  |  |
| No | 871 (52.1%) | 446 (49.6%) | 425 (55.0%) |
| Yes | 801 (47.9%) | 453 (50.4%) | 348 (45.0%) |
| **Among those with active commuting** |  |  |  |
| Average annual km/week | 38.4 (37.8) | 40.4 (39.5) | 35.7 (35.3) |
| Average annual MET-h/week | 18.8 (25.0) | 21.3 (27.4) | 15.6 (20.9) |
| **Commute greenness** |  |  |  |
| Less than 50% | 933 (55.8%) | 496 (55.2%) | 437 (56.5%) |
| 50% or more | 739 (44.2%) | 403 (44.8%) | 336 (43.5%) |
| **Mental health** |  |  |  |
| Anxiety symptoms |  |  |  |
| Minimal | 1392 (83.2%) | 711 (79.1%) | 680 (88.0%) |
| Mild | 222 (13.3%) | 147 (16.4%) | 75 (9.7%) |
| Moderate | 51 (3.1%) | 35 (3.9%) | 16 (2.1%) |
| Severe | 8 (0.5%) | 6 (0.7%) | 2 (0.3%) |
| Mental wellbeing score (0–100) | 65.3 (17.0) | 64.9 (17.3) | 65.8 (16.6) |

Values are means (standard deviations) for continuous variables and number of observations (percentages) for categorical/binary variables.

Primary commuting modes during wintertime and summertime, and transitions between seasons, are presented in Supplemental Figure 3. In short, the prevalence of active commuting as the primary mode increased from wintertime (26%) to summertime (39%). Especially cycling increased its popularity: cycling was the primary mode only for 8% of the participants during wintertime, but for 23% during summertime. More details of commute mode transitions are available in Supplemental Table 1.

### Active commuting and mental health

We observed no statistically significant dose-response relationships between active commuting and anxiety symptoms or mental wellbeing assessed in *km/week* in the minimally (Supplemental Figure 4), or fully adjusted models (Figure 2). The commute greenness interaction terms were also non-significant in all models (P_interactions_>0.167).

**Figure 2.**
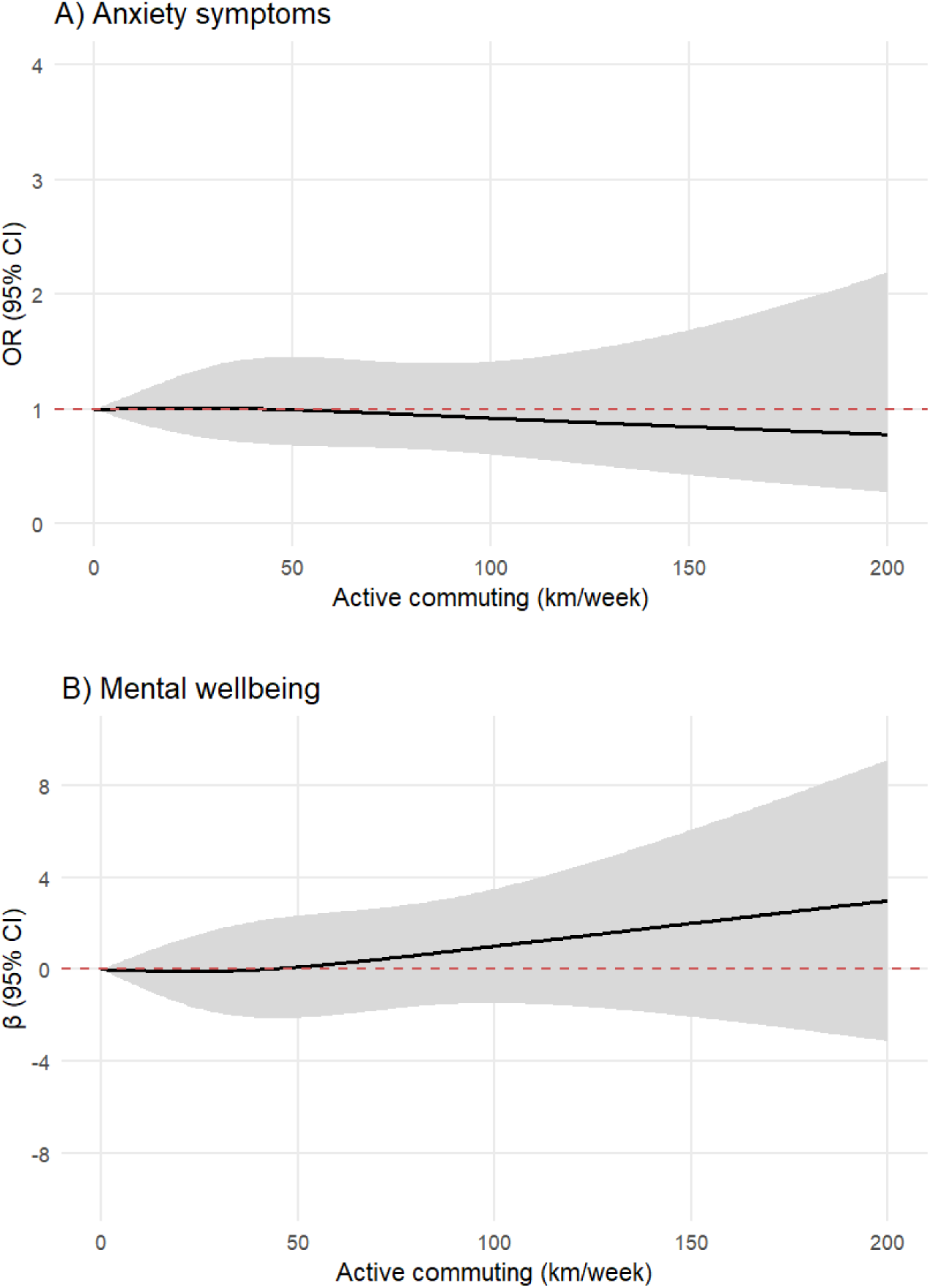
Dose-response relationship between average annual active commuting (km/week) and anxiety symptoms (panel A), and mental wellbeing (panel B). Values are odds ratios (OR) for anxiety symptoms and unstandardized betas (β) for mental wellbeing (in black) with 95% Confidence intervals (gray). No active commuting (0 km/week) was the reference. Models were adjusted with age, gender, commute greenness, education, income satisfaction, relationship status, smoking, alcohol use, leisure-time physical activity, and occupational physical strain.

We neither observed statistically significant dose-response relationships when active commuting was defined as *MET-h/week* in the minimally adjusted models (Supplemental Figure 5) or in the fully adjusted models (Figure 3). The commute greenness interaction terms were again non-significant (all P_intercation_>0.123).

**Figure 3.**
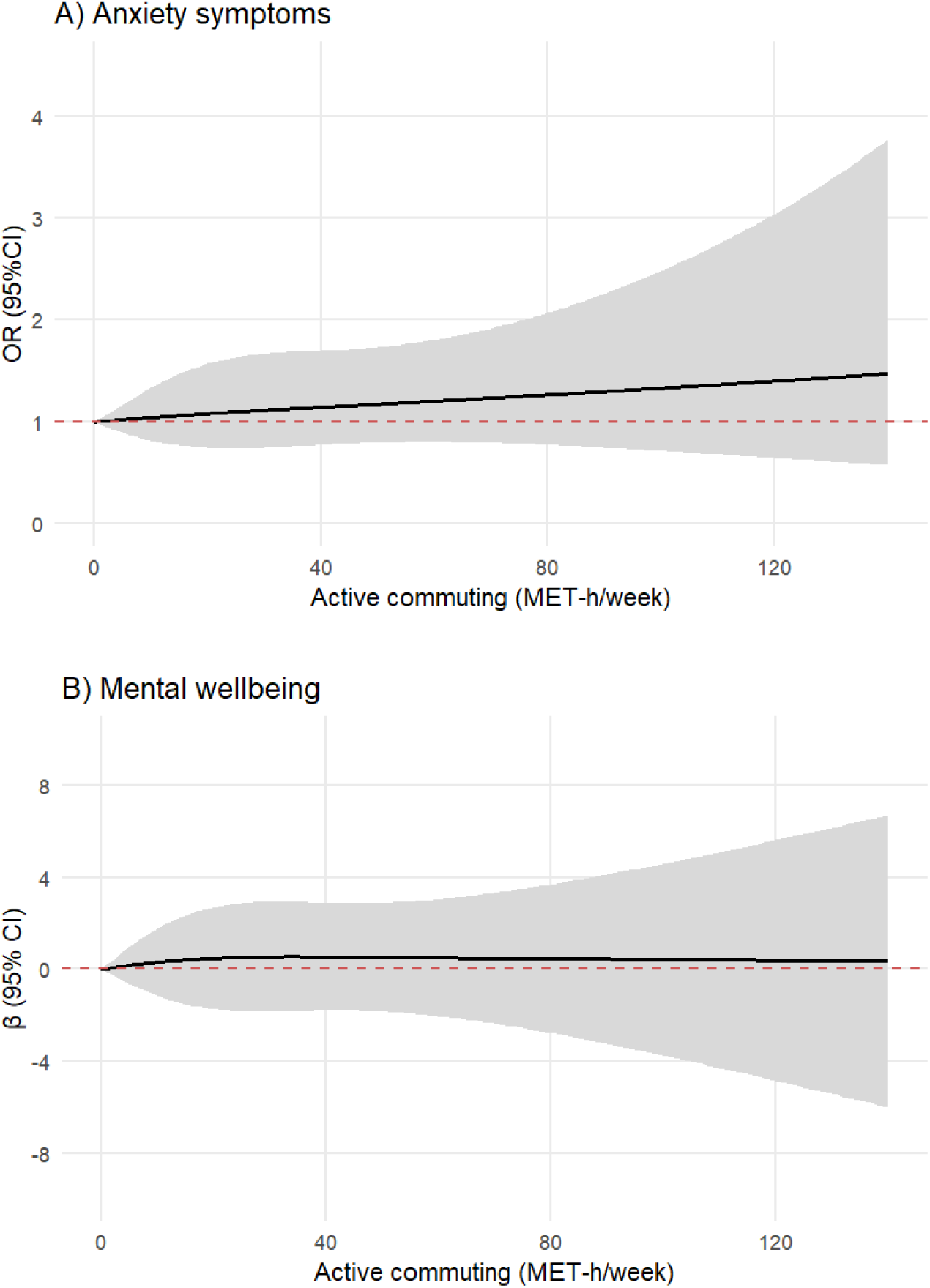
Dose-response relationship between average annual active commuting (MET-h/week), anxiety symptoms (Panel A), and mental wellbeing (Panel B). Values are odds ratios (OR) for anxiety symptoms and unstandardized betas (β) for mental wellbeing (in black) with 95% Confidence intervals (gray). No active commuting (0 MET-h/week) was the reference. Models were adjusted with age, gender, commute greenness, education, income satisfaction, relationship status, smoking, alcohol use, leisure-time physical activity, and occupational physical strain.

#### Sensitivity analyses

Adding BMI to the models did not affect the observed associations (Supplemental Figures 6 and 7). Analyses with only summertime commuting also provided similar results as the annual averages (Supplemental Figures 8 and 9).

## Discussion

In this cross-sectional study of employed adults living in the ten largest cities in Finland, we did not observe dose–response relationship between active commuting and anxiety symptoms or mental wellbeing, whether quantified as annual km/week or MET-h/week. While perceived commute greenness was positively correlated with active commuting, it did not moderate its association with mental health.

Overall, our findings are in line with most of the previous literature.^22,28^ Active commuting is a subcategory of active travel, characterized by its regularity and its potential to accumulate substantial amounts of weekly physical activity. Despite its somatic benefits, it is also characterized by ambivalent motivation, where commuting is undertaken by choice or necessity, and by generally lower affect associated with commuting itself.^45,46^ The discrepancy between somatic and mental health benefits of active commuting may be explained by the relatively consistent cardiometabolic effects of physical activity across settings, whereas mental health outcomes are more strongly influenced by the context in which the activity occurs.^47^ Context-specific influences on mental health include the type/mode of physical activity (“what you do”), the domain (“when and why you do it”), the physical environment (“where you do it”), the social environment (“who you do it with”) and the mode of delivery (“how it is delivered”), all of which are intertwined with individual preferences.^48^ The potential of active commuting to enhance mental health may be inherently limited by factors that typically link physical activity to better mental wellbeing, such as positive social interaction, a sense of autonomy, and enjoyment.^48^

Environmental modifications, such as greener routes, are promising candidates to enhance mental health during active commute.^19^ In our study, greener commutes were correlated with higher levels of active commuting, being consistent with previous research that green spaces along the travel routes may facilitate selection of active modes.^49^ However, despite the psychophysiological potential of nature environments, no effect moderation could be demonstrated in the present study. We also anticipated a dose-response pattern where longer commutes may be associated with more adverse mental health outcomes, potentially due to lesser enjoyment.^46^ However, no indication of dose-response relationships were observed in this sample, potentially due to bias toward highly educated and generally healthy participants in the study.

### Strengths and limitations

Key strengths of this study include the use of recent data from multiple Finnish cities, availability of both summertime and wintertime commute metrics and various individual-level covariates, as well as the application of flexible dose–response modelling to a topic with substantial societal relevance. We quantified active commuting using both distance-based and energy expenditure–based metrics, enhancing comparability with public health guidelines and population impact tools.

Several limitations should be also acknowledged. All measures were self-reported, which may introduce recall, social desirability bias and misclassification. The measure for green commute was coarse, and other methods might be more adequate, e.g. actual routes and Geographic Information System-based greenness data. We also acknowledge that there are several factors related to active commuting, such as infrastructure, and caring responsibilities, which were not addressed in this study. Furthermore, the survey was sent to a population-based sample of Finnish adults (N=20,000); however, the analytical sample was relatively highly educated and physically active which may limit the generalizability of the findings to a broader employed population. In addition, participants reported rather good mental health, which reduced variability in the mental health outcomes.

## Conclusion

In this study, we observed no dose-response relationship between active commuting and anxiety symptoms or mental wellbeing. The associations were not moderated by perceived commute greenness. Future research is needed to develop modifications to active commuting that promote psychological well-being and, consequently, strengthen the status of active commuting within the planetary health framework also from a mental health perspective.

## Perspective

Planetary health perspective aims to better understand and address the ways in which human and natural systems affect one another’s health and wellbeing.^1^ To mitigate climate change, legal institutions, such as the European Union, have set considerable reduction targets for greenhouse gas emission, and Finland’s national Climate Act states that carbon neutrality should be achieved by 2035 at the latest.^6^ According to recent national statistics, traffic contributes 23% of all greenhouse gas emissions in Finland, of which 52% are attributable to private cars.^50^ Many of the driven trips could be replaced with active modes as national surveys indicate that 22% of the trips <1km, and about half of 1–3 km trips are travelled with private cars.^7^ From the planetary health perspective, current and previous studies suggest that active commuting is not strongly associated with either benefits or harms to mental health. Active commuting seems, however, to support better cardiometabolic health and functional capacity, providing support for active commuting in the planetary health framework.

## Statements

### Data availability statement

Data is available upon a reasonable request from TL.

### Ethics approval statement

The data collection was approved by THL Ethics Working group (THL/706/6.02.01/2023) and conducted in accordance with the Declaration of Helsinki.

### Participant consent statement

Respondents were informed about the use and handling of data in the covering letter, and they have accepted the use of data for scientific research by returning the survey.

