## Supplemental for "Active commuting, anxiety symptoms and mental wellbeing: a dose-response study"

### Supplemental Material

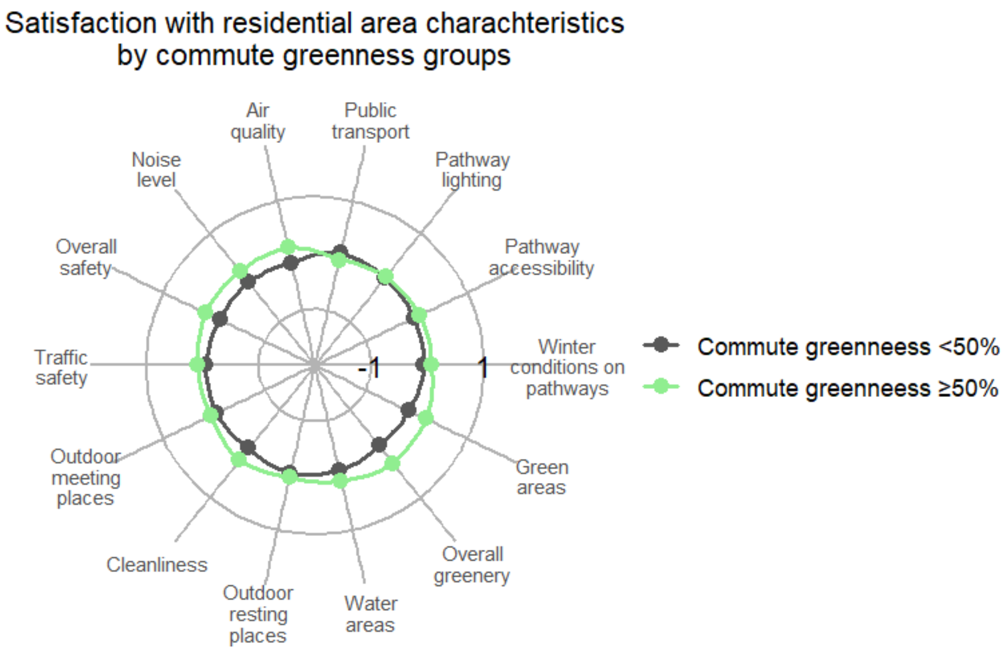


Supplemental Figure 1. Satisfaction with residential area characteristics by commuting greenness categories. Green line ≥50% greenness during commute; grey line, <50% greenness during commute. Values are Z-scores where 0 indicates sample average, -1 towards lower satisfaction and +1 towards higher satisfaction.

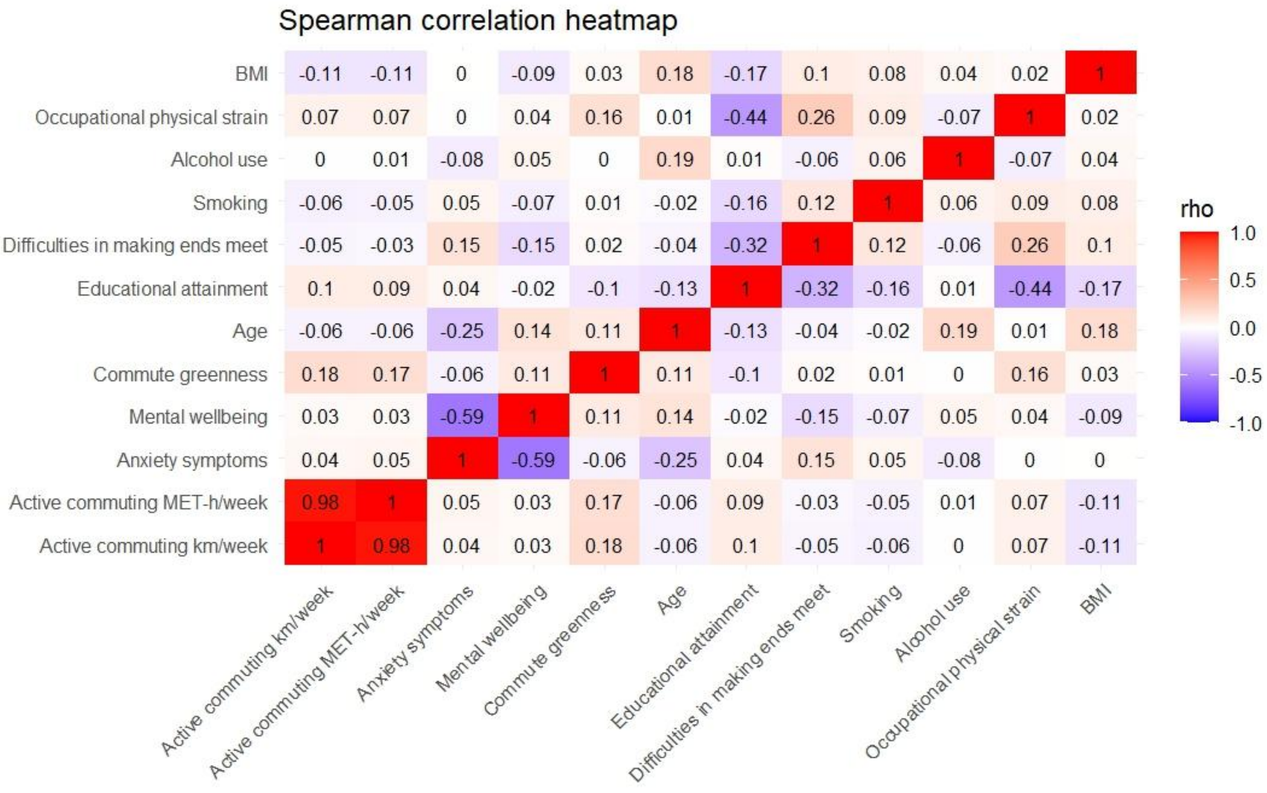


Supplemental Figure 2. Spearman correlations between continuous active commuting and outcome variables, and categorical commute greenness and covariates.

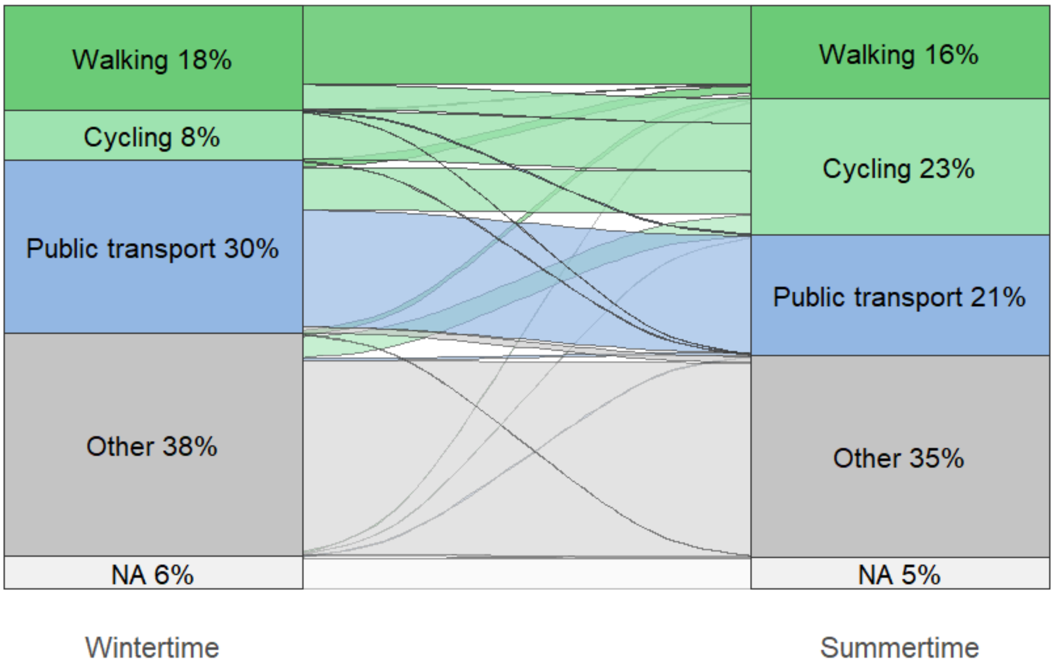


Supplemental Figure 3. Primary commuting modes during wintertime and summertime. Stratums indicate the prevalences of primary commuting modes during wintertime and summertime, and flows the transitions between seasons. In case the participants reported to use two or more modes with similar frequencies, the primary mode was walking > cycling > public transport > other. Other includes cars (driving or as a passenger), motorcycles, e-scooters and other vehicles. NA, no reported commutes.

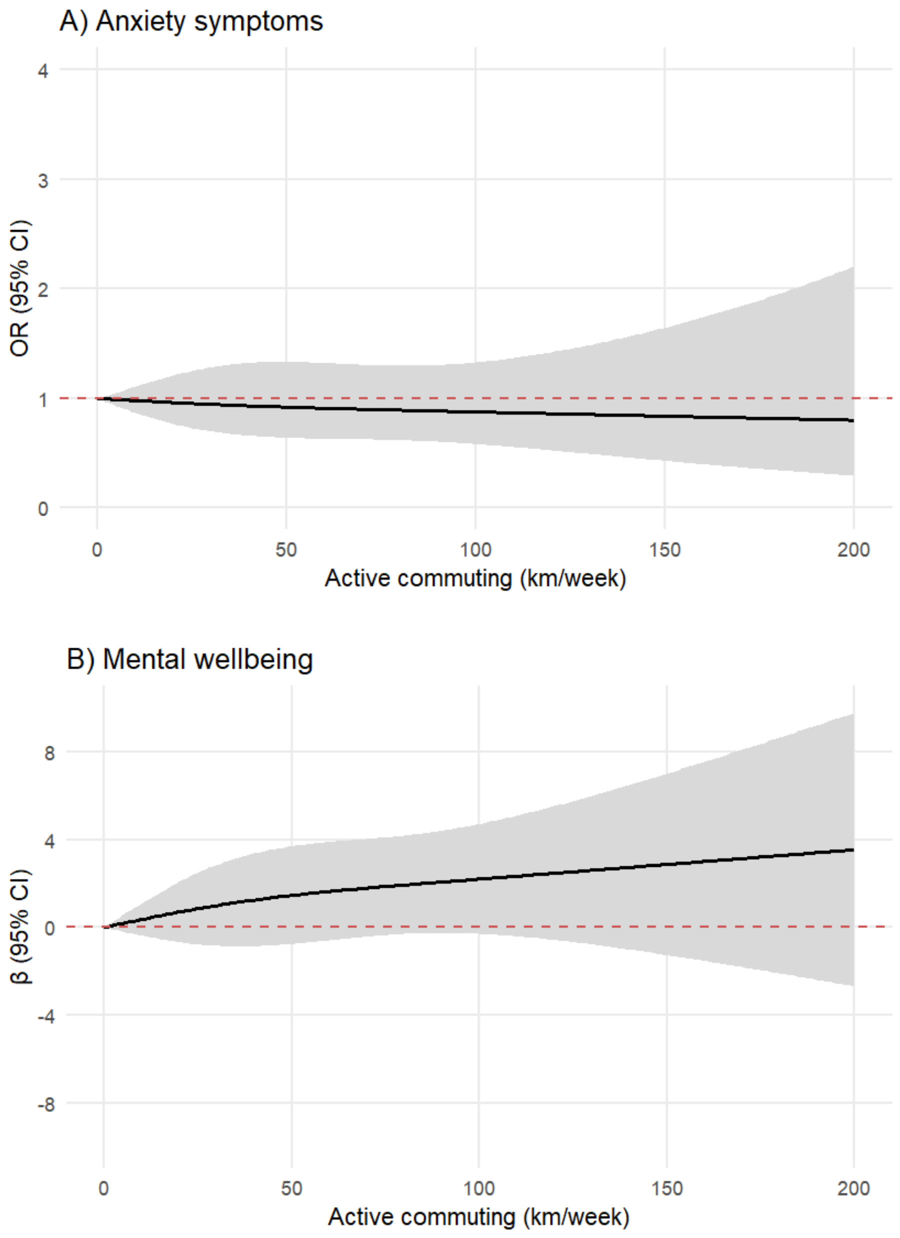


Supplemental Figure 4. Dose-response relationship between average annual active commuting (km/week) and anxiety symptoms (Panel A), and mental wellbeing (Panel B) following minimal adjustment. Values are odds ratios (OR) for anxiety symptoms and unstandardized betas (β) for mental wellbeing (in black) with 95% Confidence intervals (gray). No active commuting (0 km/week) was the reference. Models were adjusted with age and gender.

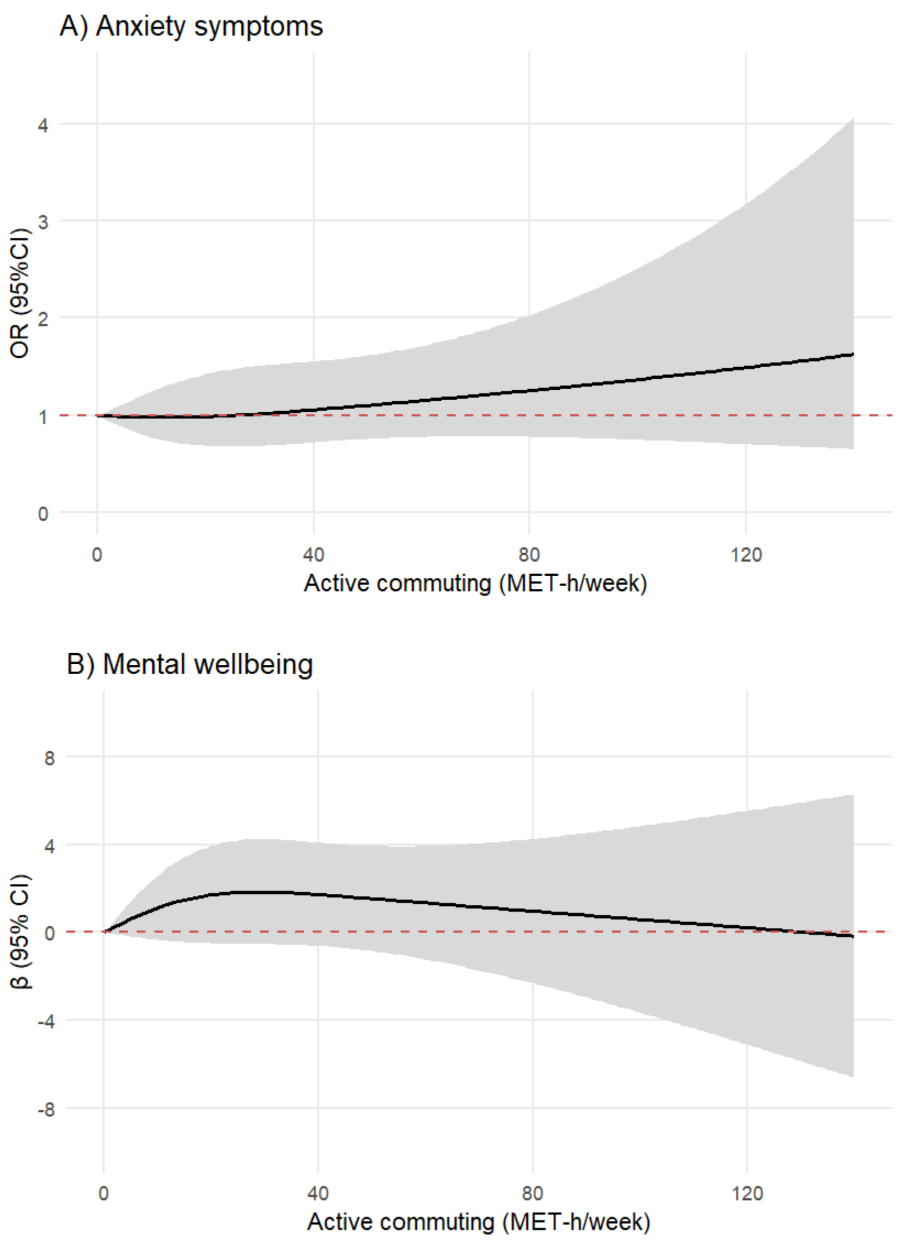


Supplemental Figure 5. Dose-response relationship between average annual active commuting (MET-h/week) and anxiety symptoms (Panel A), and mental wellbeing (Panel B) following minimal adjustment. Values are odds ratios (OR) for anxiety symptoms and unstandardized betas (β) for mental wellbeing (in black) with 95% Confidence intervals (gray). No active commuting (0 MET-h/week) was the reference. Models were adjusted with age and gender.

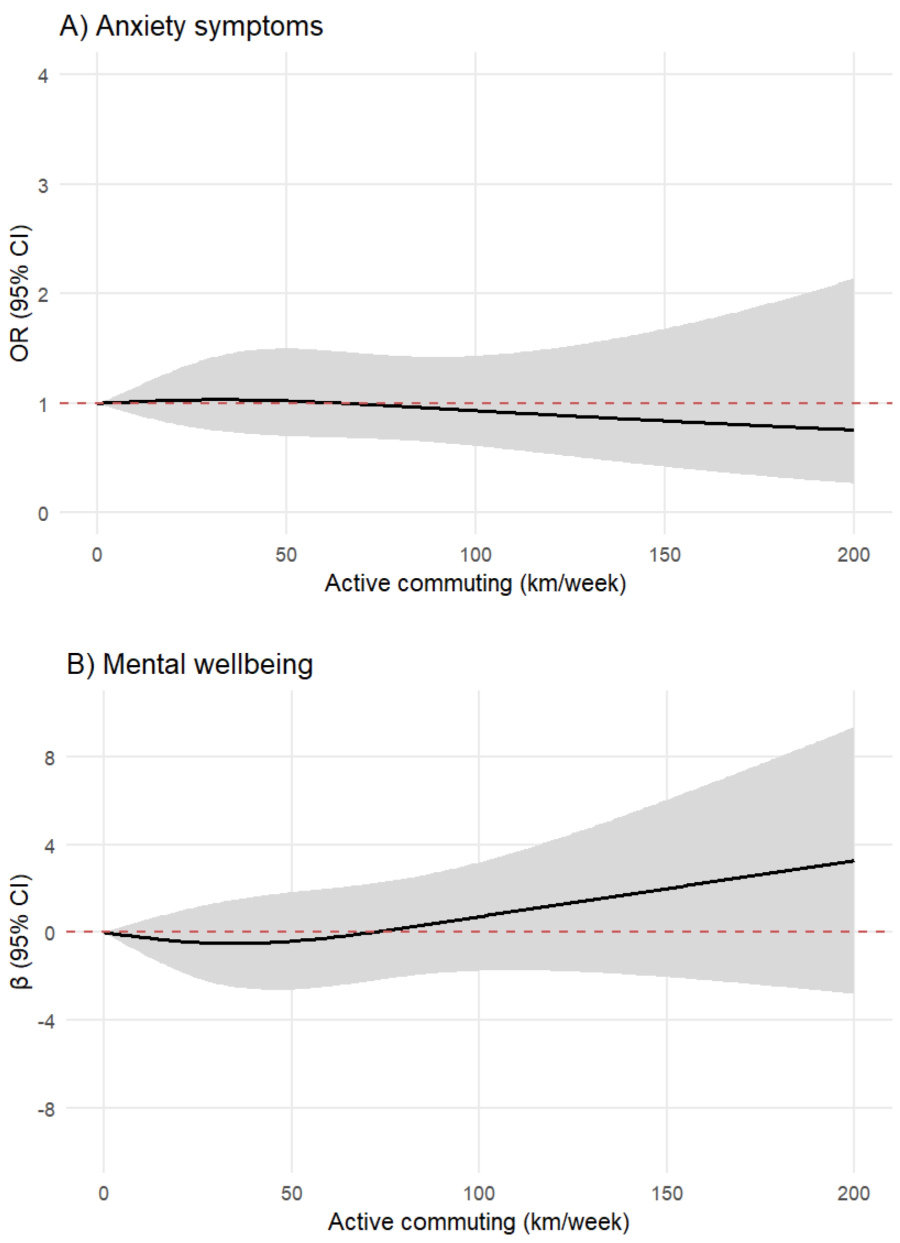


Supplemental Figure 6. Dose-response relationship between average annual active commuting (km/week) and anxiety symptoms (Panel A), and mental wellbeing (Panel B) following additional adjustment for BMI. Values are odds ratios (OR) for anxiety symptoms and unstandardized betas (β) for mental wellbeing (in black) with 95% Confidence intervals (gray). No active commuting (0 km/week) was the reference. Models were adjusted with age, gender, commute greenness, education, income satisfaction, relationship status, smoking, alcohol use, leisure-time physical activity, occupational physical strain, and BMI.

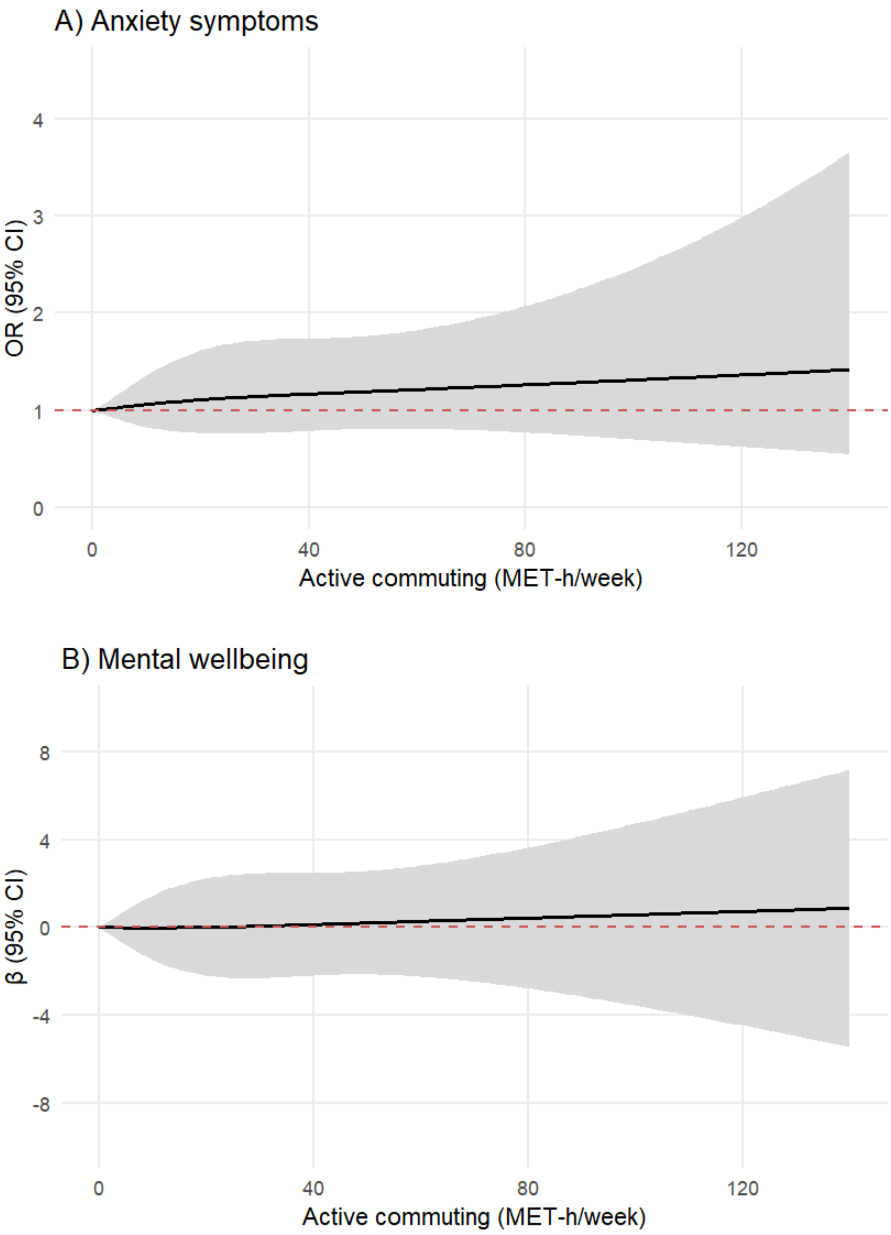


Supplemental Figure 7. Dose-response relationship between average annual active commuting (MET-h/week) and anxiety symptoms (Panel A), and mental wellbeing (Panel B) following additional adjustment for BMI. Values are odds ratios (OR) for anxiety symptoms and unstandardized betas (β) for mental wellbeing (in black) with 95% Confidence intervals (gray). No active commuting (0 MET-h/week) was the reference. Models were adjusted with age, gender, commute greenness, education, income satisfaction, relationship status, smoking, alcohol use, leisure-time physical activity, occupational physical strain, and BMI.

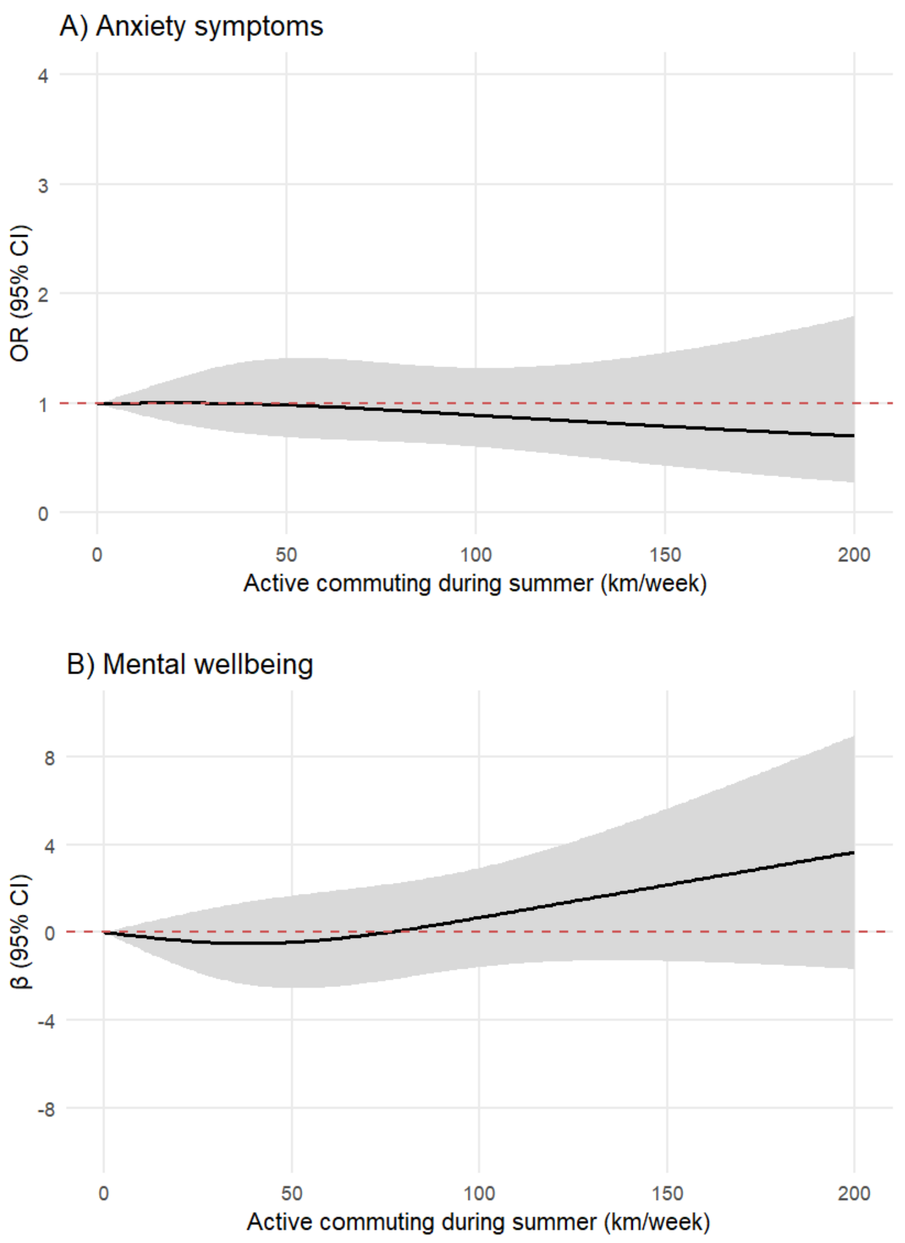


Supplemental Figure 8. Dose-response relationship between average active commuting during summer (km/week) and anxiety symptoms (Panel A), and mental wellbeing (Panel B). Values are odds ratios (OR) for anxiety symptoms and unstandardized betas (β) for mental wellbeing (in black) with 95% Confidence intervals (gray). No active commuting (0 km/week) was the reference. Models were adjusted with age, gender, commute greenness, education, income satisfaction, relationship status, smoking, alcohol use, leisure-time physical activity, and occupational physical strain.

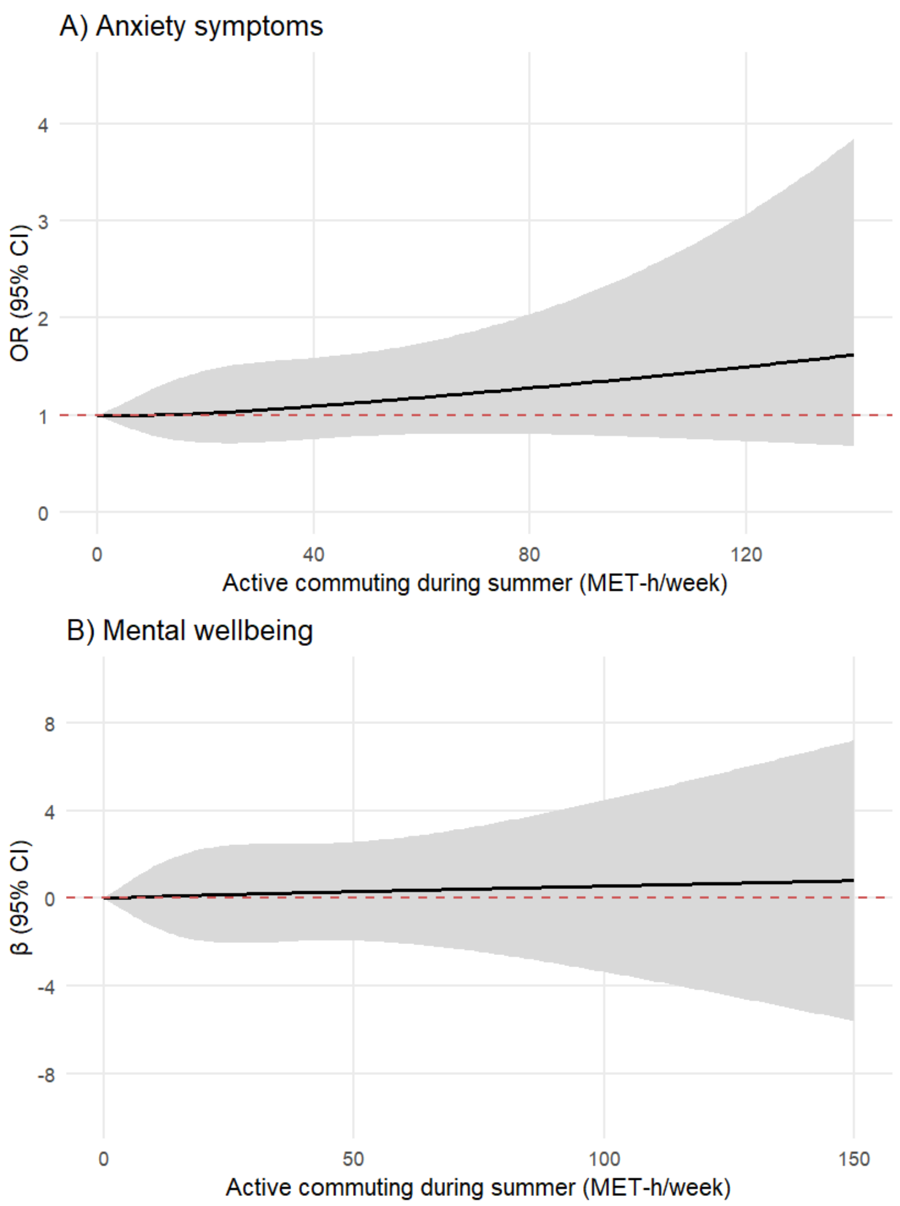


Supplemental Figure 9. Dose-response relationship between active commuting during summer (MET-h/week) and anxiety symptoms (Panel A), and mental wellbeing (Panel B). Values are odds ratios (OR) for anxiety symptoms and unstandardized betas (β) for mental wellbeing (in black) with 95% Confidence intervals (gray). No active commuting (0 MET-h/week) was the reference. Models were adjusted with age, gender, commute greenness, education, income satisfaction, relationship status, smoking, alcohol use, leisure-time physical activity, and occupational physical strain.

Supplemental Table 1. Primary commuting modes and their transitions between wintertime and summertime.

| **Wintertime** | **Summertime** | **n** | **%** |
|---|---|---|---|
| Walking | Walking | 225 | 13.5 |
| Walking | Cycling | 70 | 4.2 |
| Walking | Public transport | 3 | 0.2 |
| Walking | Other | 1 | 0.1 |
| Walking | NA | 0 | 0.0 |
| Cycling | Walking | 2 | 0.1 |
| Cycling | Cycling | 138 | 8.3 |
| Cycling | Public transport | 0 | 0.0 |
| Cycling | Other | 2 | 0.1 |
| Cycling | NA | 0 | 0.0 |
| Public transport | Walking | 22 | 1.3 |
| Public transport | Cycling | 122 | 7.3 |
| Public transport | Public transport | 334 | 20.0 |
| Public transport | Other | 17 | 1.0 |
| Public transport | NA | 1 | 0.1 |
| Other | Walking | 14 | 0.8 |
| Other | Cycling | 60 | 3.6 |
| Other | Public transport | 6 | 0.4 |
| Other | Other | 556 | 33.3 |
| Other | NA | 4 | 0.2 |
| NA | Walking | 3 | 0.2 |
| NA | Cycling | 1 | 0.1 |
| NA | Public transport | 3 | 0.2 |
| NA | Other | 2 | 0.1 |
| NA | NA | 86 | 5.1 |

NA, No reported commuting modes; Other includes cars (driving or as a passenger), motorcycles, e-scooters and other vehicles.
